# Drivers underpinning the malignant transformation of giant cell tumour of bone

**DOI:** 10.1101/2020.06.14.20129809

**Authors:** Matthew W. Fittall, Peter Ellery, Iben Lyskjær, Patrick Lombard, Jannat Ijaz, Anna-Christina Strobl, Dahmane Oukrif, Maxime Tarabichi, Martin Sill, Christian Koelsche, Jonas Demeulemeester, Grace Collord, Roberto Tirabosco, Fernanda Amary, Peter J. Campbell, Stefan Pfister, David T.W. Jones, Nischalan Pillay, Peter Van Loo, Sam Behjati, Adrienne M. Flanagan

## Abstract

The rare benign giant cell tumour of bone (GCTB) is defined by an almost unique G34W oncohistone mutation in the H3.3 histone gene. Here we reveal the genomic and methylation patterns underlying the rare clinical phenomena of benign metastases and malignant transformation of GCTB.

---

Giant cell tumour of bone (GCTB) is a locally destructive benign tumour, prone to local recurrence. They present predominantly at the site of the mature epiphysis/epimetaphysis of the long bones, particularly the distal femur and proximal tibia in the 3^rd^ and 4^th^ decades of life^1^. GCTB is defined by a near universal (96%) pathognomonic H3F3A G34W missense mutation^2-6^. Two unexplained phenomena in GCTB are of particular interest, namely that lung metastases occur despite the absence of malignant histological features in either the primary or metastatic lesions^7^ and secondly that the characteristic H3.3 mutation is occasionally found in primary malignant bone tumours which often share features with conventional GCTB^4,8^. We set out to explore the genomic events underlying these phenomena using whole genome sequencing and genome-wide methylation profiling using methylation array and whole genome bisulfite sequencing.

We started our investigation by performing whole genome sequencing (WGS) on seven primary malignant bone tumours possessing an H3.3 mutation, one case of metastatic GCTB, and nine conventional GCTB for which we had frozen tissue with corresponding germline DNA from blood samples (**Supplementary Table 1)**. We used the analysis pipeline of the *Cancer Genome Project* to generate catalogues of somatic mutations, indels, structural variants and copy number changes and a previously reported strategy to identify putative drivers (**Methods**)^9^.

Benign GCTB genomically resemble other benign mesenchymal tumours (**Figure 1a**). They possess few somatic changes of any type and no plausible driver mutations other than the canonical H3.3 mutation (medians: 640 substitutions, 43 indels, 7 structural variants; **Figure 1a and Supplementary Figure 1**). In contrast, we found that malignant bone tumours with the same H3.3 mutation possess genomic features resembling osteosarcoma. They possessed an increased burden of somatic variants, and broadly divided into two groups: 3/7 tumours exhibited a modest increase in mutations (medians: 1815 substitutions, 86 indels, 21 structural variants) and the remaining four possessed a greater mutation burden (medians: 4177 substitutions, 205 indels, 108 structural variants; Figure 1a and **Supplementary Figure 1**).

**Figure 1.**
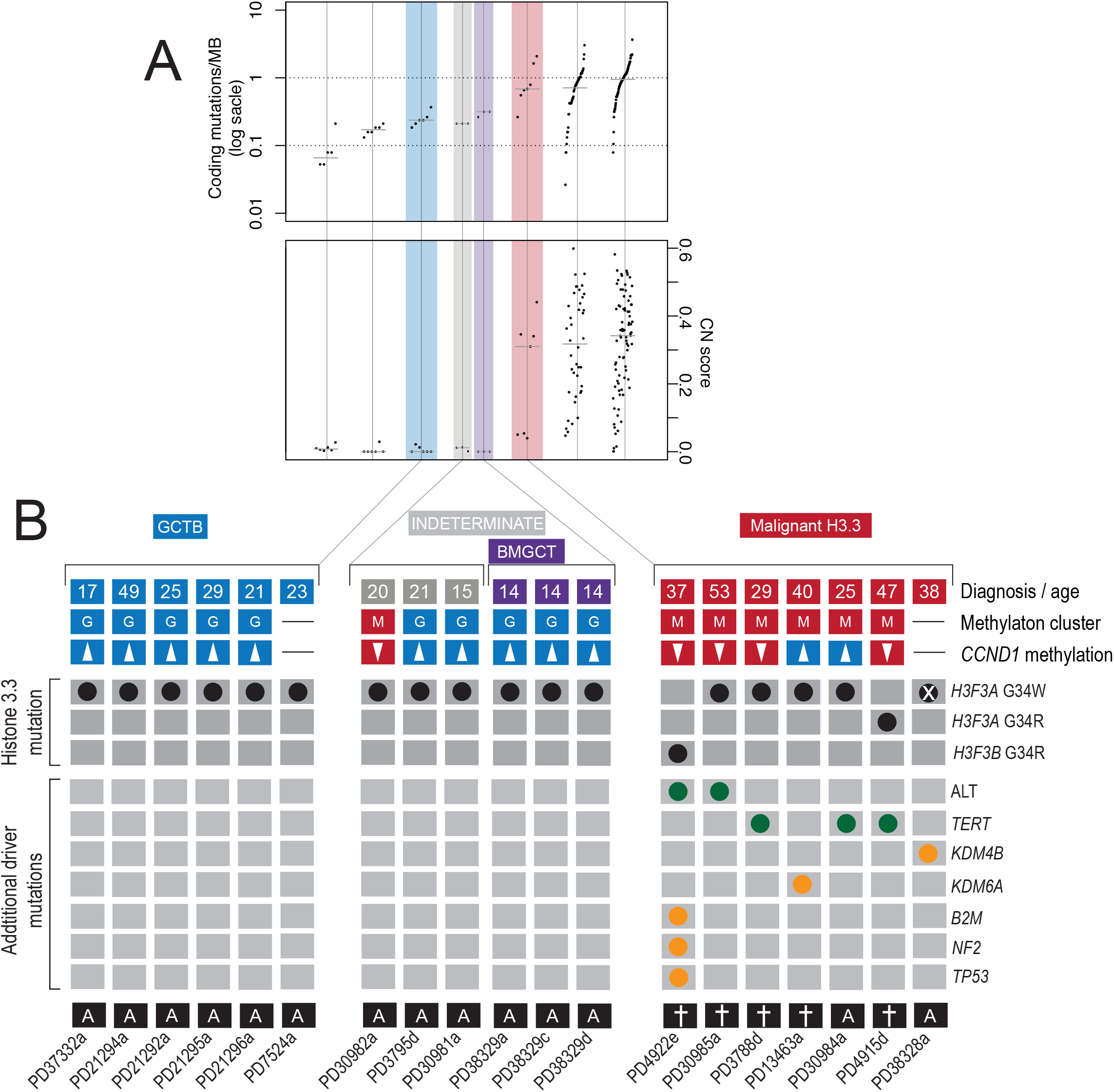
Landscape of H3.3 mutant tumours. a) Mutational burden of samples in comparison with selected other mesenchymal tumours: osteoblastoma^23^, chondroblastoma^5^, chondrosarcoma^24^ (*exome data only; SVs not shown), and osteosarcoma^9^. b) The genomic and methylation classification of sequenced tumours. From top to bottom: clinical diagnoses and age, unsupervised methylation cluster assignment, *CCND1* promoter methylation status (hypermethylation is defined as a mean *CCND1* promoter methylation beta value >0.2 and a tileplot of curated drivers, clinical outcomes are shown underneath (more detailed clinical outcomes are shown in **Supplementary Table 1**). Note sample PD38328a had undergone deletion of the H3F3A locus, which had been present on the pre-resection biopsy (**Supplementary Figure 11**)

Unlike osteosarcoma, malignant H3.3-mutated bone tumours are enriched with mutations suggesting telomere dysfunction. Two tumours had mono-allelic G>A mutations 124bp upstream of the *TERT* transcription start site, reported to increase promoter binding^10^. Another sample, PD3788d, had a complex rearrangement event, resembling chromothripsis, encompassing *TERT*, resulting in the juxtaposition of the gene *MEGF10* with the *TERT* promoter (**Supplementary Figure 2**). *MEGF10* is reported to be under the control of a super-enhancer in the dbSUPER database^11^. Two other malignant samples, PD4922e and PD30985a, were identified as having markedly elongated telomeres (**Figure 1b, Supplementary Figure 1** and **Methods**), a finding consistent with the Alternative Lengthening of Telomeres (ALT) mechanism, which is usually mutually exclusive with *TERT* alteration^12,13^. In keeping with this pattern of ALT, recently reported in other sarcoma types^14^, these tumours possessed highly rearranged genomes, the telomeres of which comprised conventional (‘TTAGGG’) repeats and had undergone loss of heterozygosity at the *RB1* locus (**Supplementary Figure 3**). In total, 5/7 malignant tumours had evidence of a *TERT*-mutated phenotype. In contrast, only *TERT* amplifications have previously been reported in osteosarcoma^9^. The remaining two malignant tumours both harbored biallelic losses of an additional histone lysine demethylase, *KDM4B* or *KDM5A* (**Supplementary Figure 3**). All malignant tumours had thus acquired at least one additional driver mutation in addition to the G34W.

**Figure 2.**
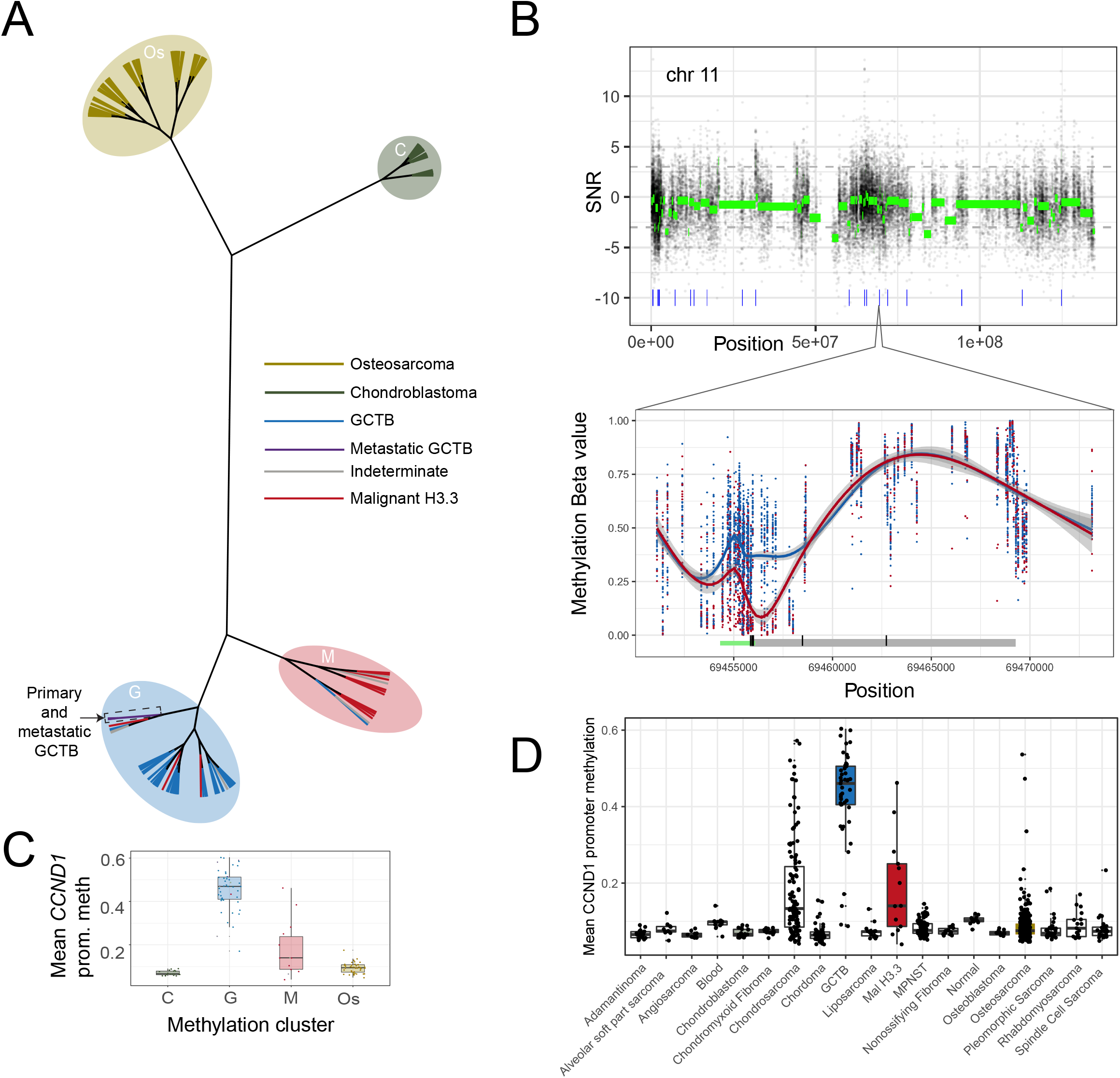
Methylation changes of H3.3 mutant tumours. a) Hierarchical (unrooted) clustering of tumours. Leaves are coloured by diagnosis and the methylation clusters annotated with shaded ovals. b) Analysis of methylation differences between malignant (“M”) and benign (“G”) tumours (n=12 and 40 respectively) across chromosome 11 (upper) and across *CCND1* (lower). Raw (black) and segmented (green) signal-noise-ratio (SNR; >0 shows greater methylation in malignant tumours) are plotted. Blue ticks in the upper plot represent DMRs. In the lower plot raw methylation beta values across *CCND1* are shown for each sample. The underlying schematic represents the *CCND1* gene body (grey) and the predicted promoter (green). c) Mean *CCND1* promoter across the clustered samples from a) and d) a variety of other tissues.

The degree of aneuploidy observed in 3/7 malignant tumours, against a backdrop of almost ubiquitously diploid GCTBs (**Supplementary Figure 4**), allowed the ordering and timing, in real-time, of the most significant mutational events. In all three cases (PD4922e, PD30985a, and PD3788d), whole genome duplication (WGD) had occurred in adulthood, but several years prior to diagnosis. Chromothripsis had occurred subsequent to this. In 2/3 samples (PD4922e, PD30985a), with informative data, the H3.3 histone mutation had also been duplicated, demonstrating its occurrence as an earlier mutational event prior to WGD (**Supplementary Figure 5**). This is consistent with the progression of these malignant tumours from GCTBs.

We next investigated the ‘benign metastasising GCTB’^1^. In contrast to malignant tumours, the morphology of both the metastases and primary lesion (PD38329a/c/d) was that of a conventional GCTB which was reflected in the low mutational burden and the absence of additional driver mutations (**Figure 1b and Supplementary Figure 1**). Leveraging the independent sampling across these three tumour samples increased the power to define the clonality of mutations. Clonal mutations are those found in all tumours cells whereas those in only a fraction of cells are considered subclonal. The primary tumour (PD38329a) and the two metastases (PD38329c and PD38329d) each possessed a group of private mutations to only that tumour sample. Furthermore, one cluster of mutations (**Supplementary Figure 6)** was found to be common but subclonal in all samples. This suggests that both metastases were seeded by at least one cell possessing those mutations and at least one cell that did not, a process known as polyclonal seeding.

To explore the epigenetic differences between malignant and benign H3.3-mutated bone tumours we collected additional tumours for DNA methylation array analysis (**Methods**). This collection (n = 121), included some of the sequenced samples, osteosarcomas without H3.3 mutations, and chondroblastomas, a benign tumour with an alternative H3.3 mutation, *H3F3B*:p.K27M (**Supplementary Table 2**). Unsupervised clustering based on the most variable methylation probes recapitulated the diagnostic groups (**Figure 2a and Supplementary Figure 7**). Furthermore, while closely related to conventional GCTB, the malignant H3.3-mutant tumours formed a distinct clade. The benign metastasising samples clustered with the benign GCTB group.

To unpick the methylation differences underlying the separate clustering of the benign and malignant H3.3 mutated tumours, differentially methylated regions (DMRs) were identified. This revealed focal changes in a small number of specific gene promoters (**Methods**). Of 74 DMRs identified, 56 were located around gene transcriptions start sites (**Supplementary Figure 8**). The most statistically significant DMR was also the only one identified in a plausible cancer driver gene, *CCND1*, which encodes Cyclin D1. Differential methylation spanned a promoter region of 1500bp either side of the transcription start site, a finding validated by bisulfite sequencing (**Supplementary Figure 9**). Comparing the mean methylation level across this promoter region between different bone and soft tissue tumour types revealed that hypermethylation at this site is specific to GCT (**Figure 2d**). Malignant histone-mutated tumours and chondrosarcomas were the only tumour types with a similar degree of *CCND1* promoter methylation. *CCND1* promoter methylation was concordant with unsupervised methylation clustering groups (**Figure 1** and **Figure 2**). Beyond this, methylation differences were enriched at non-enhancer intergenic sites, however those that affected genes did not consistently alter gene pathways. At a broader scale part of the cluster of histone genes on chromosome 6 was focally hypermethylated in malignant tumours, suggesting additional epigenetic driver events (Supplementary **Figure 10**).

Using comprehensive genomic and methylation profiling, we report the driver events associated with malignant or metastatic progression of GCTBs. Malignant H3.3 mutated tumours are characterized by a methylation profile that is related but distinct from conventional GCTB. Histone mutation predates the development of aneuploidy in malignant tumours, which still occurs some years prior to diagnosis. Malignant progression requires additional genetic mutations endowing either telomere or epigenetic dysfunction, and possible additional epigenetic changes altering clusters of histone genes. This combination of genomic and epigenomic findings could potentially distinguish benign from malignant GCTs, thereby predicting aggressive behavior in challenging diagnostic cases (**Figure 1b** and **Supplementary Table 3**): it also permits malignant GCTB to be classified on a molecular basis distinguishing it from other primary malignant bone tumours. The absence of additional genetic events in metastatic, but histologically benign GCTB, and the presence of polyclonal seeding supports the longstanding hypothesis that they represent a thrombotic event.

## Methods

### Patient samples

Patients provided their written and informed consent to provide samples for this study via the UCL Biobank for Health and Disease, based at the Royal National Orthopaedic Hospital. This was approved by the National Research Ethics Service (NRES) Committee Yorkshire & The Humber – Leeds East (15/YH/0311). DNA was extracted from areas of fresh frozen tissue selected by bone pathologists (A.M.F./R.T./F.A./P.E.). Matched normal DNA was acquired from blood samples.

### SNP and Methylation Array

SNP analysis was performed using the Illumina HumanOmni2.5 BeadChip. DNA Methylation analysis was performed on either Illumina Infinium HumanMethylation450 or MethylationEPIC arrays. Pre-processing and quality control were performed (**Supplementary Methods**).

### Array analysis

SNP array copy number profiles were produced using ASCAT (v2.5.1). Methylation array-based copy number profiles were generated using the conumee package (v1.18.0) and a bespoke adaptation of the principles utilised by ASCAT (**Supplementary Methods**). Unsupervised clustering was performed by hierarchical clustering using the 5,000 most variable probes across samples after scrutiny for batch effects (**Supplementary Methods**). Differentially methylated probes and regions were detected using the ChAMP package (v2.14.0). Gene set enrichment analysis (GSEA) was performed using an adapted approach from the ebBayes function in the ChAMP package^15^. Bespoke analysis for regional differences in methylation were performed using Circular Binary Segmentation (CBS) functions from the DNACopy package (v1.58.0) based on a signal-noise ratio for each methylation probe (**Supplementary Methods**).

### Whole genome bisulfite sequencing

Whole genome bisulfite sequencing was performed on seven of the samples: PD30981a, PD30982a, PD30984a, PD30985a, PD3788d, PD3795d and PD4915d. Oxidative bisulfite conversion and library preparation was done using the Cambridge Epigenetix Truemethyl Whole Genome kit following manufacturer’s instructions. The efficiency of bisulfite treatment was determined using control probes; 90.6% of 5-methylcytosine remaining unconverted and 100% of unmethylated cytosines were converted into thymines. Hydroxymethylation was not detected using this kit as 91.7% of 5-hydroxymethyl cytosine were converted to thymine. Libraries were sequenced on an llumina HiSeqX using a 150bp paired end run. Both mapping of reads to GRCh37 and methylation calling was done using Bismark (https://github.com/FelixKrueger/Bismark).

### Whole genome sequencing

For whole genome sequencing the Illumina (Illumina, Chesterford, UK) no-PCR library protocol was used to construct short insert 500□bp libraries, prepare flowcells and generate clusters. Whole genome sequencing was performed using the Illumina HiSeq 2000 or 2500 platform, using 100 bp paired-end libraries. Samples PD37332, PD3788, PD3795, PD38328, PD38329, PD4915, PD4922 were sequenced using the XTen platform using 150 bp paired-end libraries.

### Variant detection, validation and analysis

The Cancer Genome Project (Wellcome Trust Sanger Institute) variant calling pipeline was used to call somatic mutations (versions as below). All variant calling algorithms were used with standard settings with limited post-processing filtering and variants were analysed using a previously documented strategy^9^ (**Supplementary Methods**). Variants were considered as potential drivers if they presented in established cancer genes (Chapter 3 COSMIC v82, Chapter 4 COSMIC v85). Tumour suppressor coding variants were considered if they were annotated as functionally deleterious by the VAGrENT algorithm (http://cancerit.github.io/VAGrENT/). Disruptive rearrangements or homozygous deletions of tumour suppressors were also considered. Additionally, homozygous deletions were required to be focal (<1□Mb in size). Mutations in oncogenes were considered driver events if they were located at previously reported hot spots (point mutations) or amplified the intact gene. Amplifications also had to be focal (<1□Mb) and result in at least 5 copies in diploid genomes, or 4 copies more than the modal major copy number in genome duplicated samples.

### Copy number scoring

A sample was considered Whole Genome Duplicated (WGD) when the modal total copy number was >2. The baseline total copy number was considered as 4 for WGD samples and 2 for others. Autosomal copy number segments were then scored as the difference from this baseline; no difference (0), total copy number of 0 (homozygous deletion, 2), total copy number >= 3 + baseline (amplification, 2), other score not equal to baseline (1). Scores were normalised relative to the length of the chromosome, summed and then divided by the theoretical maximum (44). Aneuploidy was validated using image cytometry on nuclear extracts from formalin fixed tissue sections (**Supplementary Methods**).

### Mutation clustering, phylogenetic reconstruction and timing analyses

The algorithm DPClust (v2.2.6) and its pre-processing pipeline (v1.0.8) were used to cluster mutations according to fraction of cancer cells (Cancer Cell Fraction, CCF) in which they were found, as described previously^20^ (**Supplementary Methods**). Phylogenetic reconstruction was performed using the pigeon-hole principle^20^. In brief, subclones were designated to be nested within a clone or another subclone if their combined CCF exceeded that of their parent.

Initial timing analysis required the transformation of individual mutation allele frequencies into mutation copy number. This was performed using the equation:

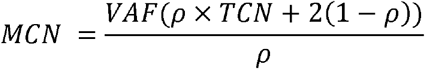

MCN is Mutation Copy Number, ρ is the sample purity, TCN is the local total copy number.

For WGD timing, deamination (clock-like, C>T mutations at CpG dinucelotides) mutations were selected from regions of balanced gain (2+2) or LOH (2+0). A probabilistic approach to WGD timing was taken with confidence intervals generated by bootstrapping sampling iterations of the underlying mutations (**Supplementary Methods**).

## Data availability

DKFZ raw methylation array data (IDAT files) were acquired from the authors of Koelsche *et al*, the data having been acquired as described^22^.

The authors declare that all data supporting the findings of this study are available within the article and its supplementary files or from the corresponding author on reasonable request. Sequencing data have been deposited at the European Genome-Phenome Archive (http://www.ebi.ac.uk/ega/) that is hosted by the European Bioinformatics Institute.

## Data Availability

The authors declare that all data supporting the findings of this study are available within the article and its supplementary files or from the corresponding author on reasonable request.

http://www.ebi.ac.uk/ega/

## Acknowledgements

This work was supported by funding from: The Tom Prince Cancer Trust, the UK Medical Research Council grant (MR/M00094X/1), The Wellcome Trust, Skeletal Cancer Action Trust UK, the Royal National Orthopaedic Hospital NHS Trust R&D Department, the Rosetrees and Stoneygate Trusts (M46-F1), the Pathological Society of Great Britain and Ireland (PL) and the Bone Cancer Research Trust. A.M.F is a NIHR senior investigator. AMF and N.P. were supported by the National Institute for Health Research, UCLH Biomedical Research Centre and the UCL Experimental Cancer Centre. M.W.F, J.D., M.T., and P.V.L. are supported by the Francis Crick Institute, which receives its core funding from Cancer Research UK (FC001202), the UK Medical Research Council (FC001202), and the Wellcome Trust (FC001202). Personal fellowships have been granted to S.B. (Wellcome Trust Intermediate Clinical Research Fellowship, St. Baldrick’s Foundation Robert J. Arceci International Innovation Award), I.L. (Lundbeck Foundation, award 2018-3018), P.J.C. (Wellcome Trust Senior Clinical Research Fellowship), N.P. (CRUK Clinician Scientist Fellowship (grant number 18387). and E.M. (CRUK Career Development Fellow), M.W.F. (from a CRUK accelerator award C422/A21434), J.D. (Research Foundation – Flanders, FWO Postdoctoral Fellowship; European Union’s Horizon 2020 Research and Innovation programme, MSCA 703594-DECODE), M.T. (European Union’s Horizon 2020 Research and Innovation programme postdoctoral fellowship, MSCA 747852-SIOMICS). P.V.L. is a Winton Group Leader in recognition of the Winton Charitable Foundation’s support towards the establishment of The Francis Crick Institute. We are grateful to the RNOH Musculoskeletal Pathology Biobank team for consenting patients and accessing samples. We thank all the patients for participating in our research and the clinical teams involved in their care.

## Author Contributions

AMF conceived the project. M.W.F. performed the data analyses to which P.L., P.E, I.L. and N.P. contributed some preliminary analyses. J.I. performed whole genome bisulfite analysis. A-C.S. performed DNA extraction. D.O. contributed the image cytometry analysis. A.M.F., P.E., R.T. and F.A. curated and reviewed the samples, clinical data and/or provided clinical expertise. M.S., C.K., I.L. and D.T.W.J. contributed to the methylation analyses. M.T. contributed to the methylation copy number analysis and the timing analyses. J.D. contributed to discussions. A.M.F., P.V.L., and S.B. directed the research. M.W.F, P.V.L., S.B., and A.M.F. wrote the manuscript.

## Supplementary Legends

**Supplementary Table 1. Clinical table of sequenced cases**

**Supplementary Table 2. Clinical table for methylation and SNP array cases**

**Supplementary Table 3. Clinically indeterminate cases**

**Supplementary Figure 1. Telomere lengths and mutation burdens for sequenced tumours**

**Supplementary Figure 2. *TERT* rearrangement in PD3788d**

**Supplementary Figure 3. *KDM4B* homozygous deletion and *RB1* loss of heterozygosity**

**Supplementary Figure 4. SNP and Methylation array-based copy number scores**

**Supplementary Figure 5. Mutation timing in malignant samples**

**Supplementary Figure 6. Mutation clustering in metastatic samples**

**Supplementary Figure 7. Tumour methylation clustering with other malignant bone tumours**

**Supplementary Figure 8. Selected gene promoters identified as differentially methylated regions**

**Supplementary Figure 9. Whole genome bisulfite sequencing confirmation of *CCND1* differential methylation**

**Supplementary Figure 10. Differential methylation in the HIST1 cluster**

**Supplementary Figure 11. The loss of H3.3 mutation in sample PD38328a**

